# MammaTrace – a cell-free DNA methylation plasma only assay for minimal residual disease detection in breast cancer patients

**DOI:** 10.1101/2025.05.20.25328012

**Authors:** David N Buckley, Alex J Kalfa, Gerald Gooden, Juan Pablo Lewinger, Marissa Pacheco, Jessica Gayton, Darcy Spicer, John Carpten, Daphne Stewart, Heinz-Josef Lenz, Chanita Hughes Halbert, Caryn Lerman, Joyce O’Shaughnessy, Barbara Pockaj, Bodour Salhia

## Abstract

**Introduction:** Metastatic breast cancer (MBC) remains an incurable disease with a 5-year overall survival rate below 25%. Metastases often emerge from subclinical, disseminated tumor cells that persist despite systemic therapy of primary disease – referred to as minimal residual disease (MRD). Detecting MRD is critical for identifying patients at high risk of recurrence and enabling timely intervention.

**Methods:** In this study, we developed MammaTrace, a plasma-only cell-free DNA (cfDNA) methylation-based MRD assay, informed by differentially methylated regions (DMRs) identified in MBC using whole genome bisulfite sequencing. MammaTrace was evaluated in an independent longitudinal cohort of early-stage breast cancer patients treated with curative intent.

**Results:** MammaTrace achieved a sensitivity of 91% and specificity of 83%, with a median follow-up of 12.4 months. A positive MammaTrace score, indicative of MRD, preceded clinical or radiologic recurrence by a median of 457 days, providing a substantial lead time for therapeutic intervention to prevent progression to metastatic disease.

**Conclusions:** MammaTrace enables detection of minimal residual disease in breast cancer patients, offering a substantial lead time before clinical recurrence. This approach may improve risk stratification and guide early therapeutic strategies to delay or prevent metastatic progression.

## INTRODUCTION

Breast cancer is the leading cause of cancer deaths in women globally (1). While 25% of metastatic breast cancer (MBC) presents as de novo disease, the majority of MBC arises after a diagnosis of localized early-stage breast cancer (EBC) that is usually treated systemically with curative intent (2). MBC arises from minimal residual disease (MRD), also known as molecular residual disease, which is subclinical, microscopic, disseminated cancer cells persisting in the body following primary and adjuvant therapy. MRD broadly refers to the presence of residual disease or molecular signals post-treatment and is not yet standardized across cancer types or clinical contexts. While monitoring MRD is the standard of care for hematological malignancies, its application in solid tumors — including breast cancer — is still evolving, with no fixed time points or universal detection protocols. This is further reflected in the current National Comprehensive Cancer Network (NCCN) guidelines, which do not include MRD testing and surveillance. Recent advancements have established MRD surveillance as a valuable tool in monitoring treatment response, predicting relapse, and guiding therapeutic decisions in solid tumors. By identifying trace amounts of cancer cells that remain in a patient’s body after treatment, MRD surveillance can provide early indications of recurrence long before symptoms or radiographic evidence of MBC appear, allowing for timely interventions and more personalized management of cancer. Existing MRD assays that rely on early post-treatment timepoints are primarily designed to assess the immediate predictive value of MRD for informing adjuvant treatment decisions. However, these tests are unlikely to predict recurrence in patients who would relapse at a much later timepoint, which can happen in breast cancer even up to 10 years post-treatment. Notably, long-term studies evaluating the utility of commercially available MRD testing beyond early post-treatment time frames have not been conducted, leaving a critical gap in understanding how MRD may be used for extended surveillance. Despite advances in risk stratification and the deployment of increasingly effective systemic and locoregional therapies, 20-30% of EBC patients will develop metastases. There remains a need for more sensitive diagnostic assays to identify patients with MRD following primary systemic treatment who may benefit from additional treatment and enhanced surveillance. Expanding the scope of MRD testing beyond early post-treatment timepoints and integrating it into long-term disease monitoring strategies could significantly improve breast cancer outcomes.

The development of circulating tumor DNA (ctDNA) (the fraction of cell-free (cf)DNA derived from tumor cells) assays has enabled blood-based monitoring of MRD. Peripheral blood harbors tumor-derived analytes including circulating tumor cells, microRNAs, extracellular vesicles and ctDNA, which carry tumor-specific genetic and epigenetic alterations (3, 4, 5, 6, 7, 8). There are several advantages to developing liquid biopsy assays based on aberrant DNA methylation over those detecting other molecular alterations such as point mutations or serum-based protein markers (9, 10, 11, 12, 13, 14, 15). DNA methylation changes occur more frequently across the genome than single nucleotide variants or copy number alterations, making them easier to detect (13, 16). Additionally, DNA methylation assays target multiple altered regions with numerous CpG sites, allowing for a lower limit of detection compared to protein-based markers or DNA mutations (17). Furthermore, since DNA methylation is altered in tumor cells, changes can be detected even in the presence of background cfDNA derived from normal cells (12, 17).

The use of ctDNA as a biomarker for MRD is an emerging technology that has shown promise across multiple tumor types (18, 19, 20, 21). MRD detection can be broadly divided into two strategies: tumor-informed and tumor-naïve. In a tumor-informed approach, DNA from archival tumor tissue is sequenced to identify genomic alterations that can be monitored in blood using personalized assays to detect selected mutations. However, tumor-informed assays may be limited by availability of adequate tissue. Furthermore, whole exome sequencing of the primary tumor and designing a personalized assay add logistical complexity and cost to the process. In contrast, tumor-naïve (also known as ‘tumor-uninformed’ or ‘tumor-agnostic’) MRD detection strategies do not rely on knowing patient-specific tumor alterations. Tumor-naïve approaches, therefore, do not require tumor tissue nor the production of bespoke panels to evaluate the presence of MRD (18). These approaches instead detect MRD by identifying cancer-specific alterations that are common across multiple tumors for a given cancer type (22, 23). While these tend to have broader applicability, simplified workflows, and reduced costs, they have suffered from lower sensitivity and specificity compared to tumor-informed tests (22).

In the current study, we aimed to develop a test for MRD surveillance, focusing on identifying cfDNA methylation markers specifically associated with breast cancer recurrence and metastasis, rather than markers of tumor burden. In this study, we introduce MammaTrace, a novel, tumor-naïve cfDNA methylation-based plasma-only liquid biopsy assay designed for MRD surveillance and recurrence prediction. While it does not rely on prior knowledge of the primary tumor, it is uniquely informed by molecular markers associated with metastatic disease, allowing for sensitive detection across diverse stages of cancer progression. This strategic focus on metastatic-associated markers acknowledges that disseminated breast cancer cells may persist for years or decades without progressing to clinically evident metastatic disease, suggesting that not all detectable MRD predicts recurrence. Focusing on cfDNA markers associated with recurrence risk rather than tumor burden alone may therefore increase the accuracy of the assay in predicting recurrence. Furthermore, our plasma-only discovery approach captures systemic, biologically relevant cfDNA signals directly from plasma, rather than relying on tumor tissue. This design not only increases clinical applicability, particularly in settings where tumor tissue is unavailable, but also acknowledges that cfDNA methylation patterns may better represent the dynamic and complex processes involved in cancer. Moreover, whether and how mechanisms of tumor dormancy may influence MRD is also yet to be determined (24).

MammaTrace was evaluated in a longitudinal study comprised of 107 diverse EBC breast cancer patients who were treated with neoadjuvant chemotherapy followed by surgery. MammaTrace correctly identified MRD at the end of treatment in 10 of 11 EBC patients who later developed metastatic disease (sensitivity = 0.91 at 0.83 specificity), with a median lead time of 15 months before clinical recurrence. Our findings demonstrate that MammaTrace is a highly accurate, novel cfDNA methylation MRD assay that may have clinical utility in identifying EBC patients who remain at high risk of recurrence in spite of standard curative therapy.

## METHODS

### Sample Accrual

This study analyzed a total of 579 plasma samples from 331 patients, categorized into an MBC cohort and a longitudinal early-stage breast cancer (EBC) cohort (**Table S1, Figure 1A**). About 1-4ml of plasma were collected in EDTA (purple-top) tubes and plasma harvested and stored at −80C until further use. MBC patients and healthy individuals were acquired from several sources including the University of Southern California and various commercial vendors. The EBC cohort was collected by Dr. Barb Pockaj at the Mayo Clinic, Arizona between May of 2015 through October of 2019 (**Table S1**). All patients provided written informed consent. Both cohorts were comprised of Asian, African American (AA), Hispanic/Latino (HL), and non-Hispanic White (NHW) patients (**Figure 1B**). A summary of patient demographics for the MBC/Healthy and EBC cohorts can be found in **Figure 1B, Table S2,** and **Table S3**. The MBC cohort included plasma from MBC patients (N = 95), and healthy female donors (N = 110) (**Figure 1A/B, Table S2**). To ensure that epigenetic alterations were generalizable to different populations, we also included 19 samples from African American patients with triple-negative breast cancer (TNBC) across varying stages in the MBC cohort (**Figure 1B, Table S4**). The median age of MBC patients was 52.5 years (**Figure 1B**), the median age of AA TNBC patients was 53 years, and the median age of healthy donors was 52 years (**Figure 1B**).

**Figure 1:**
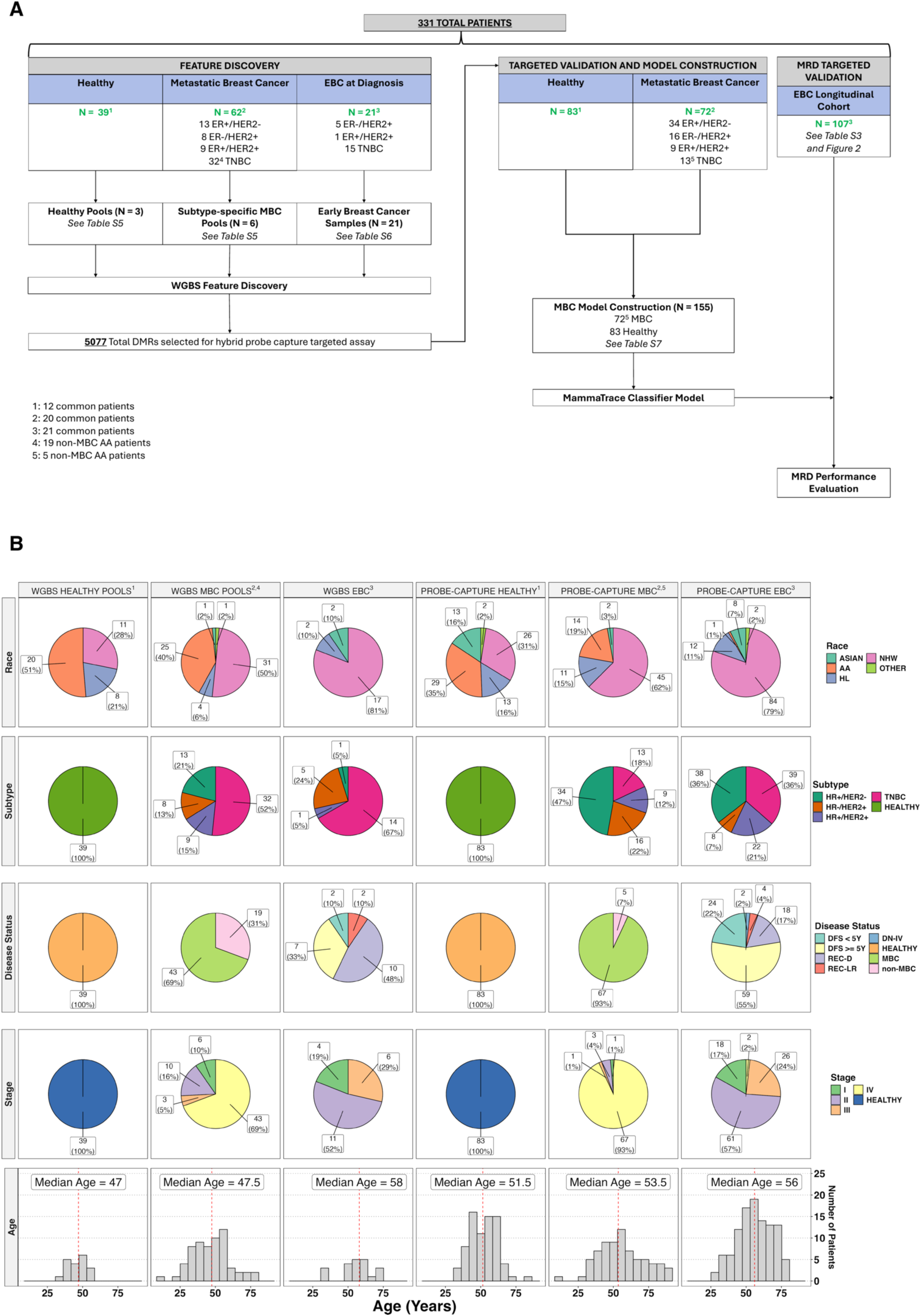
Patient sample summary and study workflow. (**A**) A total of 331 unique patientdonors were used in this study. The study used plasma from healthy individuals, patients with metastatic breast cancer (MBC) and patients with early-stage breast cancer (EBC) who were undergoing neoadjuvant chemotherapy. Patients with EBC had multiple collections over time. MBC patients were used for feature discovery of MRD markers. The number of MBC patient plasma samples used for feature discovery is shown in the left panel. The right panel shows the number of patients used for targeted validation of MRD markers and model construction. (**B**) Pie charts and histograms for demographic and clinical pathological features. Pie charts represent the breakdown of race, molecular subtype, disease status, and stage by cohort type. Histograms plot the age distribution by cohort type. Columns represent the cohort type used in the study and assay type (WGBS or hybrid probe capture). The number of samples overlapping between feature discovery and targeted validation are indicated by a superscript (1–3). The number of non-MBC African American (AA) TNBC samples are also indicated by superscripts (4–5).

For the EBC cohort, blood samples were collected from 107 patients (355 total samples) at 4 primary timepoints: at diagnosis (*baseline*), after neoadjuvant chemotherapy (*post-neo*), within 9 months post-surgery (*post-op*), and during a follow-up visit at least 9 months post-surgery (*follow-up)* (**Figure 2A, Table S1/S3**). Blood was also collected from two patients after distant recurrence (‘*post-recur*’, **Figure 2A**). Seventy-six samples were not used due to low cfDNA yields, leaving 279 samples for analysis (**Table S5**). The stage breakdown is as follows: stage I N = 18, stage II N = 61, stage III N = 26 (**Figure 1B**). Additionally, 2 patients presented with *de novo* stage IV breast cancer (DN-IV). All major molecular subtypes of breast cancer were represented: HR+/HER2− (N = 38), HR−/HER2+ (N = 8), HR+/HER2+ (N = 22), and TNBC (N = 39) (**Figure 1B, Table S3**). Patients in the EBC cohort were comprised of NHW (N = 84), 12 HL, 8 Asian, and 1 AA participants (**Figure 1B, Table S3**). Clinical outcomes for all the patients were tracked through July of 2024. Over the duration of the study, 18 patients had a distant recurrence (REC-D), 4 had a local or regional recurrence (REC-LR), and 83 patients were classified disease-free survivors (DFS) as of July of 2024 (**Figure 1B, Table S3**). The median disease-free survival time was 6.03 years from the end of primary treatment (min = 0.167 years, max = 8.38 years); 59 DFS patients were disease free for more than 5 years, 24 were disease-free for less than 5 years (**Figure 1B**). A longitudinal blood collection timeline plot is shown in **Figure 2B** and a Venn Diagram showing the overlap of patients across timepoints is shown in **Figure 2C**. Excluding the 2 *post-recur* collections, 31 patients had blood collected at 1 time point only, 18 had blood collected at 2 time points, 26 had blood collected at 3 time points, and 32 had blood collected at 4 time points (**Figure 2C**).

**Figure 2:**
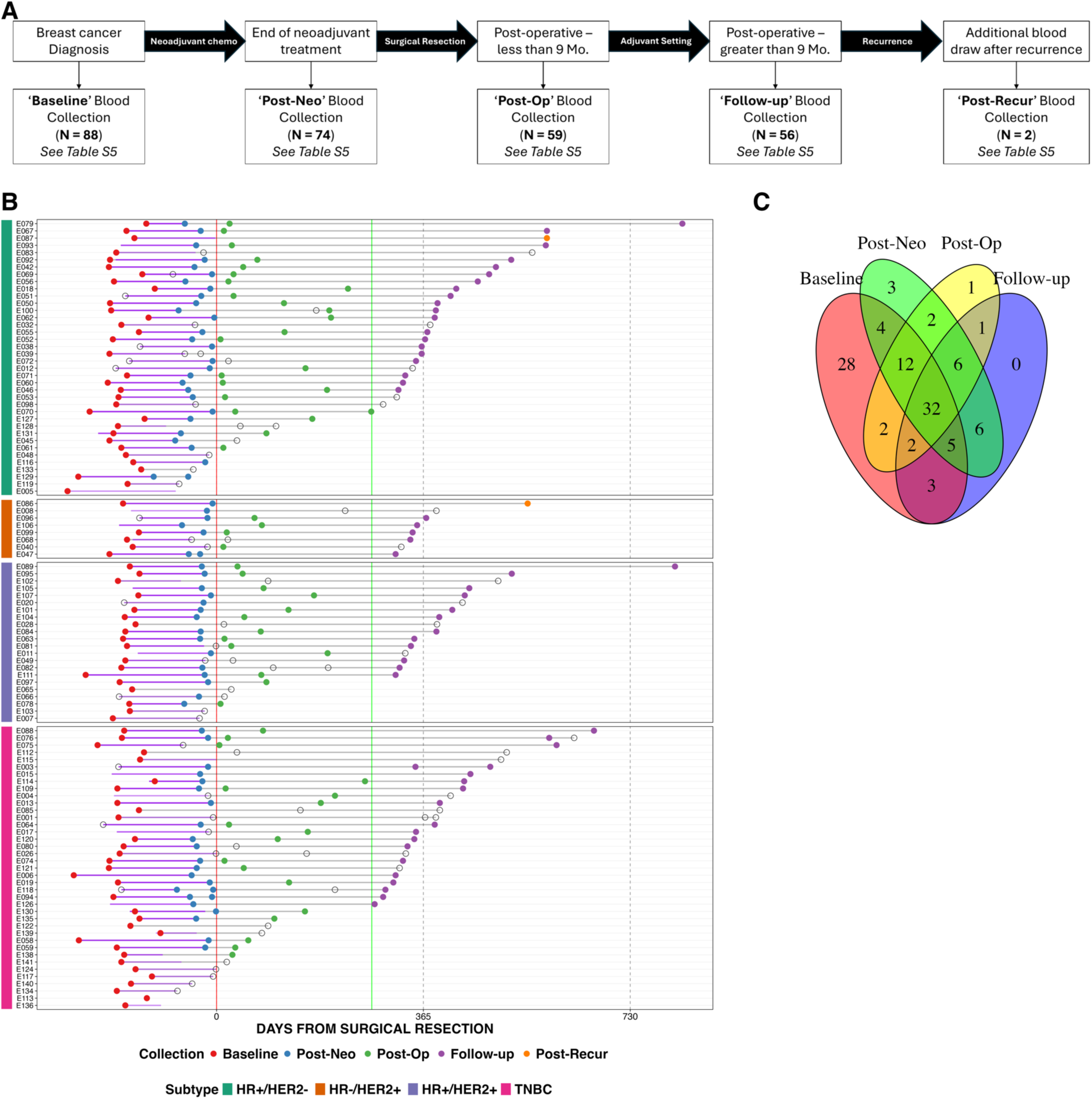
Early-Stage Breast Cancer Cohort Description. (**A**) Overview of blood collection time points for the EBC Mayo Clinic cohort. Blood was collected longitudinally from EBC patients (from 05/2015 to 10/2019) at diagnosis (*baseline*), post-neoadjuvant treatment (*post-neo*), within 9 months post-surgery (*post-op*) and at follow-up clinic visit (*follow-up*). The median follow up time was 12.4 months (range: 9 months – 27 months). A total of 107 patients with EBC were enrolled totaling 355 plasma samples collected longitudinally. Two patients had a recurrence at one of the designated blood draws and were designated “*post-recur*”. (**B**) A summary of each patient’s treatment timeline and serial blood collection grouped by breast cancer subtype (annotated in the left bar). The timelines are centered at the date of surgical resection for each patient, which is represented by the red vertical line (time = 0). Each colored dot represents one blood collection – the color denotes the timepoint of collection (baseline, post-neo, post-op, follow-up, and post-recur). The green vertical line represents the follow-up timepoint for the study. Open circles indicate samples that were collected but excluded from analysis due to QC failure. Samples not collected were not shown. (**C**) Not all patients had blood collections at every timepoint. The Venn diagram illustrates the number of patients with blood collections at every timepoint and their overlap. *Post-recur* collections (n = 2) were excluded from the diagram.

### Whole genome bisulfite Sequencing and Differential Methylation analysis

To investigate cfDNA methylation changes in MBC, we obtained 62 breast cancer plasma samples and generated subtype-specific cfDNA pools (**Figure 1A**). DNA was extracted using the Magmax Cell-Free DNA Isolation Kit from Applied Biosystems. For WGBS cfDNA subtype-specific pools were created by combining equal amounts of cfDNA per sample. Bisulfite conversion was done using the EpiTect Fast DNA Bisulfite kit (Qiagen: cat#59824). Sequencing libraries were made with the Ovation Ultralow Methyl-Seq Library kit (Nugen: cat#0335). These pools included five subtype-specific groups: an HR+/HER2 pool (N = 13 individuals), an HR−/HER2+ pool (N = 8 individuals), an HR+/HER2+ pool (N = 9 individuals), and two triple-negative breast cancer (TNBC) pools (N = 5, 6) (**Figure 1A, Table S6**). We also generated a pool of 21 samples from AA patients with TNBC across varying stages (**Table S4**). Additionally, three control pools were prepared using cfDNA from healthy individuals consisting of two predominantly NHW and HL pools (N = 9 and 10 individuals) and one healthy African American (AA) pool (N = 20 individuals) (**Table S6**).

Next, we also performed WGBS on 21 *baseline* samples from the EBC cohort (**Figure 1A**). Specifically, we sequenced the individual *baseline* samples from patients at diagnosis before they developed recurrence (REC-D N = 10, REC-LR N = 2) and the *baseline* samples from patients at diagnosis that remained DFS for greater than 5 years (N = 9) (**Table S7**). Each cfDNA sample underwent whole genome bisulfite sequencing (WGBS) as previously described (16) and sequenced to a median depth of 22X for the individual EBC samples and 53X for MBC/healthy cfDNA pools.

Metilene and DSS were used to identify differentially methylated regions (DMRs) by comparing each MBC subtype pool with healthy control groups, and by comparing the EBC *baseline* REC-D and REC-LR samples to the DFS and healthy cfDNA pools (25, 26). The AA TNBC pool was specifically compared with an AA healthy pool consisting of cfDNA from 20 individuals. Statistically significant differentially methylated regions (DMRs) were filtered using defined thresholds for methylation difference, adjusted p-values, and minimum coverage depth to ensure robust and reproducible identification. We also identified 800 regions from MBC and baseline EBC samples that exhibited cfDNA fragment methylation patterns associated with disease state. A custom pipeline developed in the Salhia lab was used to detect and analyze clusters of nearby CpG sites genome-wide. This approach characterizes read-level methylation patterns and compares their distribution between cases and controls to prioritize regions with strong discriminatory potential. A total of 5,077 unique DMRs, from both MBC and *baseline* early-stage breast cancer samples were selected as targets for hybrid probe capture assay design.

### Hybrid probe capture

To investigate the potential for the 5077 MBC-associated DMRs as markers of MRD for EBC, we designed a custom hybridization probe capture assay, to selectively enrich each of those regions. In total, the 5077 DMRs spanned approximately 1.24 Mb and encompassed 71,118 CpG loci, which were used to design a targeted hybridization probe capture panel (Daicel Arbor Biosciences). WGBS read data from MBC and EBC WGBS samples, exhibiting a range of methylation states, were used as the reference sequence for probe design. Sequencing reads aligning to the target regions were extracted and used to guide the design of an 80-nucleotide probe set with approximately 3X tiling density. Redundant or highly similar probe sequences were filtered out to reduce overlap and ensure efficient target enrichment. The final capture panel comprised approximately 474,000 probes.

Hybridization probe capture, sequencing, alignment, and methylation calling on all samples was performed as previously described (27, 28, 29). Six nanograms of cell free DNA was concentrated to a volume of 13 ml and used as template for bisulfite conversion and library preparation using the Tecan Ultralow Methyl-Seq kit following the manufacturer’s protocol (Tecan Genomics: cat#9513-A01). Final library products were quantitated using TapeStation D1000 High Sensitivity screen tapes (Agilent: cat#5067-5584). Eight libraries were combined for targeted capture using custom myBaits by Daicel Arbor Biosciences according to manufacturer protocol. The amplification product of the first hybridization probe capture was used as template for a second round of capture/wash/amplification. Final amplification products were quantitated using TapeStation D1000 High Sensitivity screen tapes. Equimolar pools of capture products were sequenced on the NovaSeq6000 (Illumina) at the Keck Genomics Platform. For NGS quality control (QC), alignment rate was calculated by Bismark (30), and cfDNA insert size distributions were calculated by qualimap bamQC (31); all QC metrics were aggregated using MultiQC (32).

### Machine Learning

To develop a robust method for detecting MRD in breast cancer, we utilized cfDNA data by targeted hybridization probe capture sequencing. A support vector machine (SVM) binary classifier was trained using sequencing data from patients with MBC (N = 72) and healthy individuals (N = 83) (**Figure 1A, Table S8**). To enrich for ctDNA, cfDNA sequencing data were preprocessed to retain only read pairs with fragment lengths below 150 bp, a threshold previously associated with ctDNA enrichment and improved classifier performance (33, 34). Next, for each target region, we computed the proportion of unmethylated (U), mixed (X), and methylated (M) reads mapped to each region. This fragment-level “UXM” approach has demonstrated utility in cfDNA-based tissue deconvolution (35). Of the 5,077 identified DMRs, 4,049 regions passed quality control filtering and were used as features for model training. Methylation data were processed using a custom, proprietary pipeline developed in-house to optimize feature extraction and classifier performance. Model training and evaluation were conducted using five repetitions of five-fold cross-validation to ensure robust performance assessment (36). After cross-validation, a final model — referred to as the MammaTrace model — was trained using the full dataset and subsequently applied to the EBC cohort for MRD detection.

## RESULTS

### cfDNA methylation differences associated with breast cancer metastasis

To identify cfDNA methylation alterations associated with MBC for use in developing an MRD assay, we employed a two-pronged strategy. This approach aimed to identify DMRs linked to distant recurrence by analyzing both diagnostic EBC samples collected prior to recurrence and MBC samples collected after recurrence. For this, we performed WGBS on 6 cfDNA pools, including 5 MBC pools representing each molecular subtype, plus a separate AA TNBC pool. Two healthy female pools from predominantly NHW women and one pool from healthy AA women were used as controls. In total, the 6 MBC cfDNA patient pools represented 62 individuals and the 3 healthy cfDNA pools represented 39 individuals. A breakdown of the number of patients in each pool can be found in **Table S6**). DMRs were identified between subtype-specific MBC cfDNA pools and healthy controls (**Figure 1A, Table S6**). AA TNBC was compared only to the AA healthy pool to control for race-specific methylation alterations. We identified a total of 2,353 DMRs from MBC pools versus healthy control pools—specifically, HR+/HER2− N = 270 DMRs, HR−/HER2+ N = 362 DMRs, HR+/HER2+ = 507 DMRs, TNBC N = 1214 DMRs (401 TNBC DMRs were derived from the AA-specific TNBC pool). The DMRs for each subtype were largely unique (**Figure 3A**). We also examined the overlap between DMRs identified from cfDNA in predominantly NHW TN MBC and AA TN MBC patients. Only 4 DMRs overlapped between the 817 NHW TNBC-associated DMRs and the 397 AA TNBC-associated DMRs, (**Figure 3B**), however these patients were not stage-matched. These results highlight the significant impact of race on cfDNA methylation changes in TNBC and possibly other breast cancer subtypes. Hierarchical clustering and uniform manifold approximation and projection (UMAP), based on beta values in MBC DMRs, showed clear separation of the MBC sample pools from the healthy control pools (**Figure 3C/D**).

**Figure 3:**
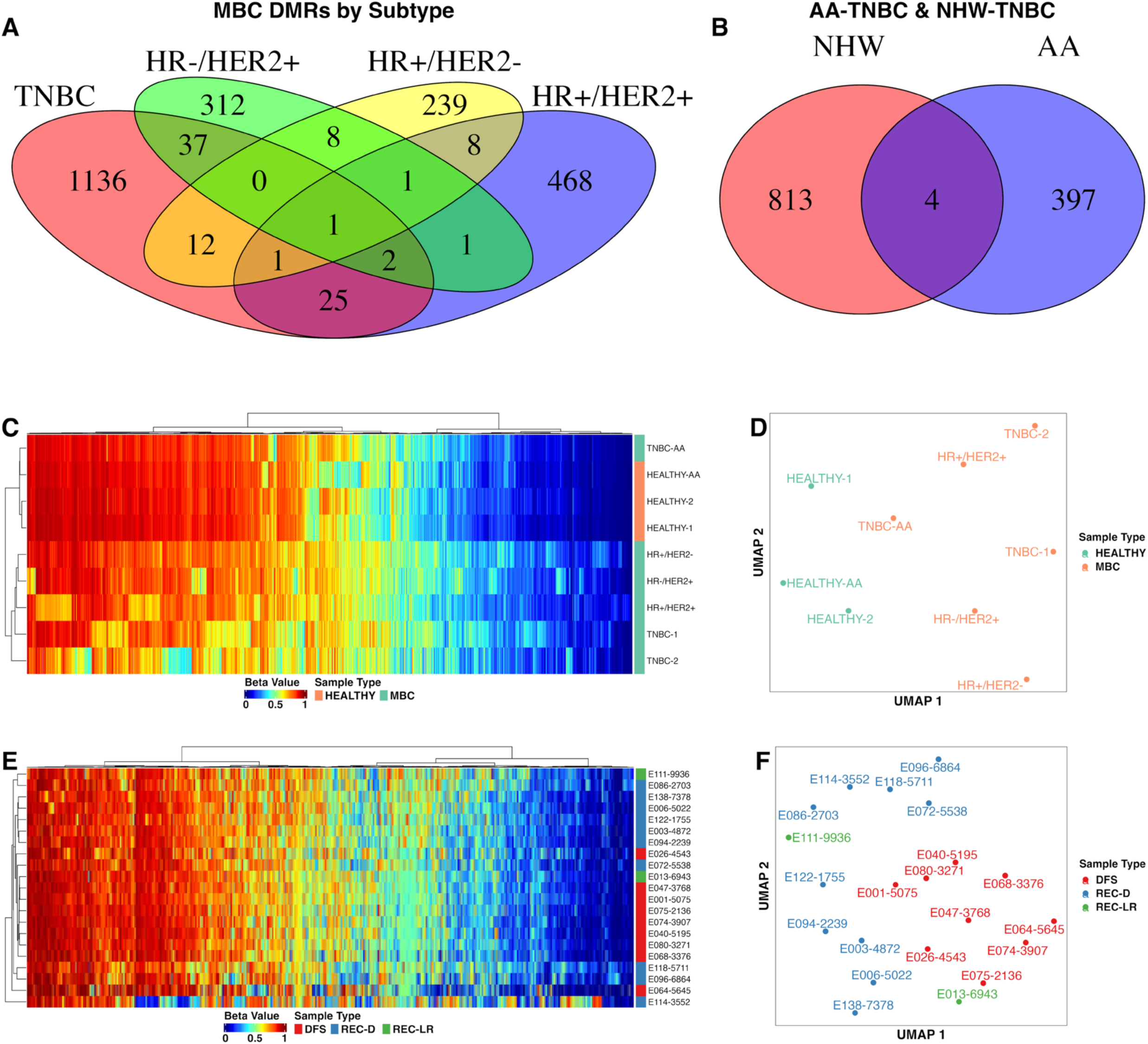
Feature discovery for MRD markers from cfDNA WGBS analysis. **(A)** Venn diagrams showing overlap of DMRs identified in subtype-specific MBC cfDNA pools. (**B**) Venn diagram showing overlap of DMRs between NHW and AA TNBC cfDNA pools. (**C/D**) Heatmap and UMAP based on mean beta values across all MBC-associated DMRs, demonstrating separation between healthy and MBC pools used for feature discovery. (**E/F**) Heatmap and UMAP based on mean beta values across all early-stage breast cancer associated DMRs, showing separation by disease status, including DFS, REC-D, REC-LR, and NDF patients.

Next, we performed WGBS on 21 *baseline* samples from the EBC cohort (**Figure 1A**). Specifically, we analyzed the *baseline* samples from patients at diagnosis before they developed distant (REC-D, N=10) or locoregional recurrence (REC-LR, N=2), and we also analyzed the *baseline* samples from patients at diagnosis who remained disease-free for greater than 5 years (N=9) (**Table S7**). These *baseline* samples comprised all major breast cancer molecular subtypes - TNBC (N = 14), HR−/HER2+ (N = 5), HR+/HER2− (N = 1) patient, and HR+/HER2+ (N = 1) (**Table S7**). We performed DMR analysis to compare both locally and distant recurrence patients to those who remained recurrence-free, and to the two non-AA healthy control pools. We found 2,338 significant DMRs in REC samples when compared to disease-free and healthy samples. Hierarchical clustering/UMAP, based on recurrence status showed a clear separation between the REC and disease-free samples (**Figure 3E/F**).

### Validation of DMRs and construction of the MammaTrace classifier for MRD in MBC samples using hybridization probe capture

In this study, we initially pooled cfDNA from MBC samples for WGBS, which—while comprehensive—is resource-intensive. Pooling offered several advantages: it reduced cost, enhanced the detection of common biologically relevant methylation changes across a patient group, improved the signal-to-noise ratio, and optimized resource use, particularly in the context of limited cfDNA analyte availability (37). We later analyzed unpooled baseline EBC samples from patients who either remained cancer-free for at least five years or eventually developed MBC. From this analysis, we prioritized 5,077 DMRs associated with metastasis, as described in the Materials and Methods.

As a first step in evaluating these regions, we developed a targeted hybridization probe capture assay based on the 5,077 DMRs to assess its ability to distinguish MBC from healthy controls. Using this probe panel, we performed hybridization capture sequencing on cfDNA from 72 MBC and 83 healthy individuals (total N = 155). The MBC samples included 34 HR+/HER2−, 16 HR−/HER2+, 9 HR+/HER2+ and 13 TNBC patients (**Figure 1B, Table S8**). Of these, 20 MBC and 12 healthy samples were included in pooled WGBS for DMR discovery (**Figure 1A**). The cfDNA validation cohort included 43 AA, 15 Asian, 24 HL, and 71 NHW individuals (**Figure 1B, Table S8**). Additionally, two samples were sequenced across seven separate runs and served as technical replicates throughout the study. These controls showed highly consistent beta values at the target loci across all runs, demonstrating excellent reproducibility and the absence of batch effects (**Figure S1**). These findings are consistent with our previous work showing that hybridization probe capture for cfDNA methylation analysis is both highly reproducible and accurate (27, 28).

To develop a classifier that distinguishes metastatic breast cancer (MBC) from healthy control cfDNA samples, we employed a repeated, nested 5-fold cross-validation strategy: an outer loop to estimate generalization performance and an inner loop to optimize hyperparameters (19, 38, 39). Each inner and each outer CV was repeated 5 times. A receiver operator characteristic (ROC) curve, based on the model’s prediction score demonstrated excellent discrimination, with a ROC area under the curve (AUC) of 0.956 (95% CI: 0.925-0.988) (**Figure 4A/B**). Performance was consistent across molecular subtypes with slightly lower performance in TNBC (AUCs: HR−/HER2+ = 0.983, HR+/HER2− = 0.972, HR+/HER2+ = 0.953, TNBC = 0.887) (**Figure 4C**). Classifier performance also remained robust across racial and ethnic groups (AUCs: AA = 0.890, Asian= 0.990, HL = 0.994, NHW = 0.960) (**Figure 4D**). A final classifier was then trained using all available MBC and healthy control samples without outer cross-validation. The output from this model, referred to as the *MammaTrace score*, was used to predict MRD status in the EBC cohort.

**Figure 4:**
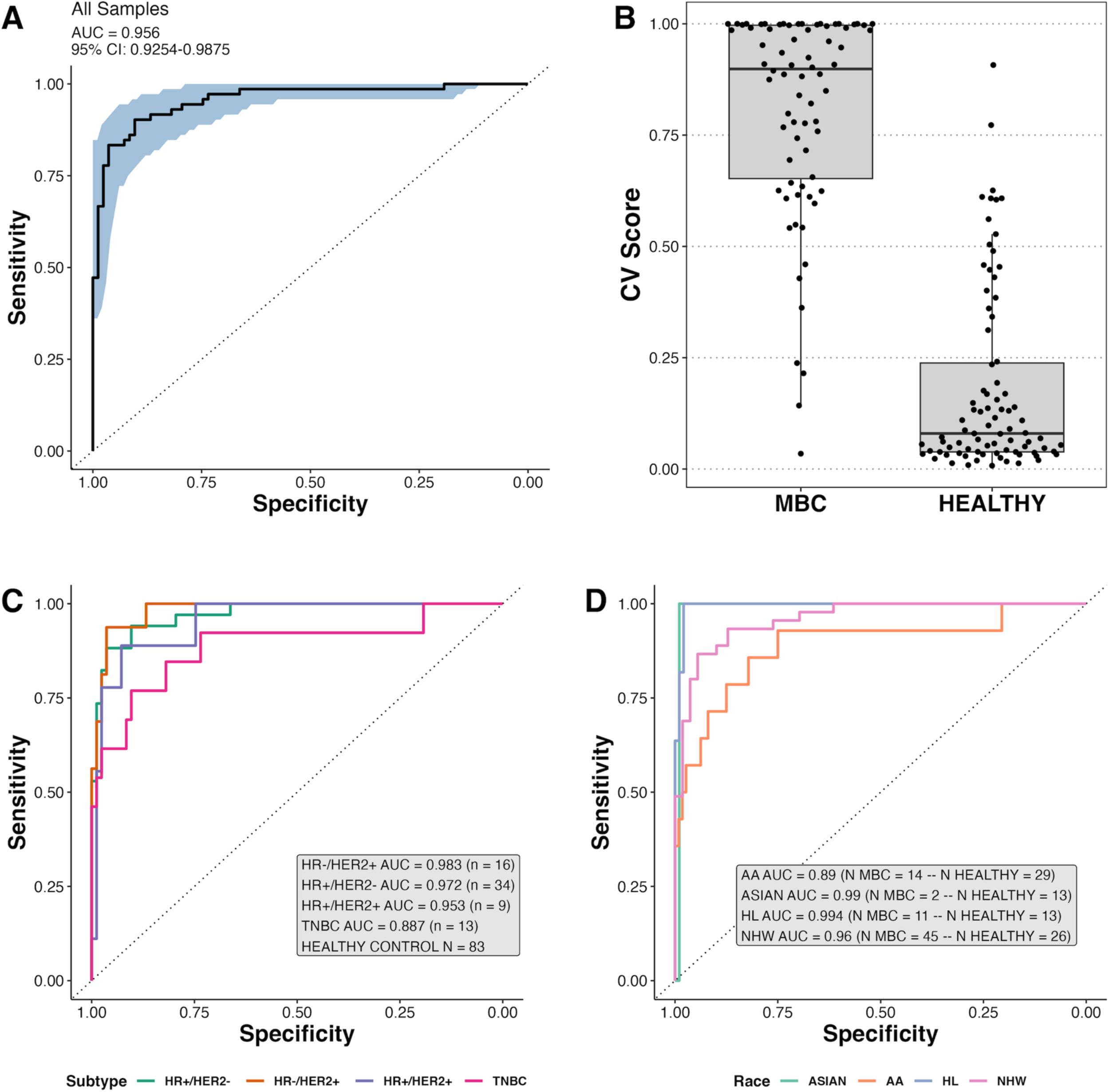
Targeted validation and construction of MammaTrace classifier. **(A)** ROC curve showing cross-validated performance of the MammaTrace model in distinguishing MBC from healthy cfDNA samples. AUC with 95% confidence interval is shown. **(B)** Boxplot of cross-validated prediction scores in an independent cohort of MBC and healthy samples not used for feature discovery. Each point represents an individual sample. **(C)** ROC curves from the same cross-validated model in (A), stratified by MBC molecular subtype. The number of MBC patients per subtype, along with the number of healthy controls, is indicated. **(D)** ROC curves from the same cross-validation, stratified by race. The number of MBC and healthy samples per group and corresponding AUC values are shown. Five non-MBC AA TNBC patients were included in this analysis.

These findings demonstrate that methylation changes associated with MBC, identified using WGBS from samples collected either prior to recurrence (baseline EBC) or after recurrence (MBC), are accurate, reproducible, and generalizable across patient subtypes and populations. This reinforces the clinical relevance of the identified DMRs and supports their use in building a classifier to distinguish MBC from healthy individuals. The resulting model lays the foundation for MRD detection in EBC patients.

### Detection of MRD in early-stage breast cancer patients using MammaTrace

To evaluate the MammaTrace assay in detecting MRD in EBC patients selected for neoadjuvant chemotherapy and surgery, we applied our custom hybrid probe capture assay to 279 cfDNA samples collected longitudinally from 107 patients with EBC (**Figure 1**, **Figure 2**). MammaTrace was initially assessed at each of the five clinical timepoints (*baseline at diagnosis*, *post-neo*, *post-op*, *follow-up* (9 months – 27 months), and *post-rec*, **Figure 2**), regardless of whether patients had collections across multiple timepoints. This initial analysis aimed to identify the most informative timepoint for MRD detection and recurrence prediction relative to patient outcomes.

In DFS patients (those without recurrence), we observed an overall increase in the MammaTrace score at the *post-neo* and *post-op* timepoints compared to *baseline*, with average scores returning to *baseline* during *follow-up*. Specifically, the median MammaTrace score increased from 0.74 at *baseline* to 0.96 *post-neo* (**Figure 5A/B**). In patients with matched samples (N=43), MammaTrace scores increased by a median ΔMammaTrace score of 0.171 at the *post-neo* timepoint (Wilcoxon’s signed rank test p-value = 0.00097) (**Figure S2**). These data suggest that cfDNA methylation patterns are altered after treatment, in contrast to ctDNA mutations, which has been shown to exhibit clearance during and after chemotherapy (40, 41, 42, 43, 44, 45). For DFS samples, MammaTrace scores subsequently decreased at the *post-op* and *follow-up* timepoints, with a median score of 0.90 at *post-op* (N = 48), and a median score of 0.75 at *follow-up* (N = 40), effectively returning to the *baseline* pre-treatment median score (**Figure 5A/B**). This trend was also observed in paired samples, with a median ΔMammaTrace score of −0.046 (*post-neo* to *post-op*, N = 42) and −0.124 from *post-op* to *follow-up* (N = 33) (Wilcoxon’s signed rank test p-values = 0.036 and 0.0082, respectively) (**Figure S2**).

**Figure 5:**
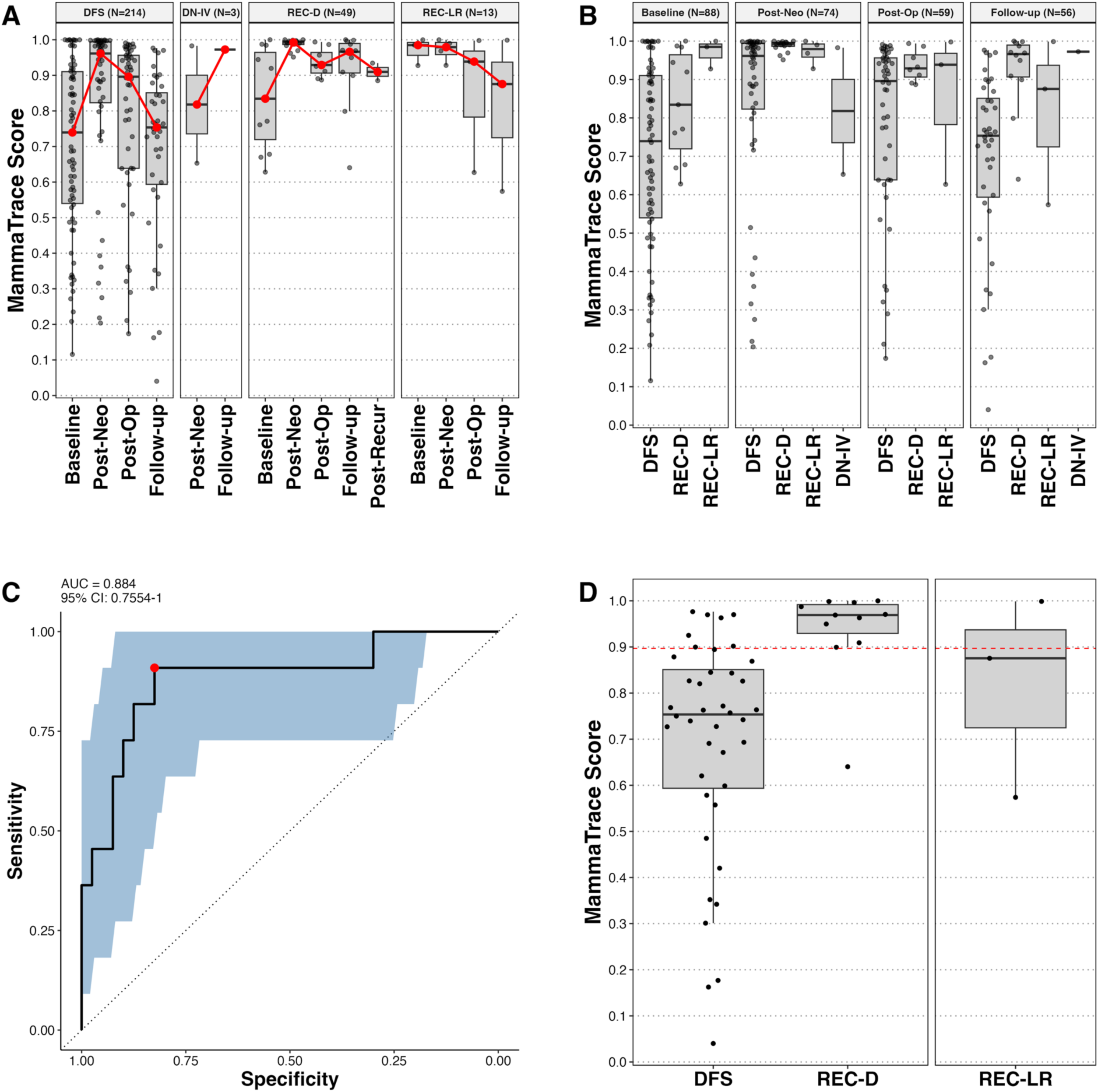
Evaluation of MRD in early-stage breast cancer using MammaTrace. **(A)** MammaTrace scores for each sample in the EBC cohort, grouped by disease status. **(B)** MammaTrace scores across treatment timepoints. **(C)** ROC curve showing the performance of MammaTrace for evaluating MRD at the follow-up timepoint. AUC and 95% confidence interval are indicated. **(D)** Boxplot showing the distribution of MammaTrace scores at the follow-up timepoint for DFS, REC-D, and REC-LR patients. The red dotted line represents the threshold for a MammaTrace-positive result, as determined by Youden’s index.

For the REC-D group (patients who developed distant recurrence), we observed a similar spike in the MammaTrace score between the *baseline* and the *post-neo* timepoints, with the median score increasing from 0.83 to 0.99 (N = 11, 16 samples, respectively) (**Figure 5A/B**). This increase was also observed in 7 paired REC-D samples, with a median ΔMammaTrace score of 0.191 (Wilcoxon’s signed rank test p-value = 0.031) (**Figure S2**). However, unlike the DFS group, the MammaTrace scores in the REC-D group did not decrease at the *post-op* and *follow-up* timepoints. Median scores remained elevated at 0.93 post-op and 0.966 at follow-up (9–27 months, N = 8 and 12 collections, respectively) (**Figure 5A/B**). In paired REC-D samples, MammaTrace scores showed a slight decrease from *post-neo* to *post-op* (median D = –0.039, *p* = 0.031) and from post-op to follow-up (median D = –0.015), though the latter was not statistically significant (*p* = 0.19) (**Figure S2**). Scores in two post-recurrence collections remained high at 0.885 and 0.934 (data not shown).

MammaTrace score dynamics were harder to assess in the DN-IV (de novo metastasis) and REC-LR (locoregional recurrence) groups due to the small number of patients (**Table S3**). In the DN-IV group, one patient had a high *follow-up* score (0.97), consistent with active disease, while the second patient lacked a viable *follow-up* sample (**Figure 2**, **Figure 5A/B**). In the REC-LR group, scores were high at *baseline* (N = 3) and *post-neo* (N = 4; scores = 0.985, 0.975), with a gradual decline at *post-op* and *follow-up* (N = 3 each; scores = 0.939, 0.875) (**Figure 5A/B**).

Next, we used ROC curve analysis to evaluate the performance of MammaTrace in distinguishing DFS from REC-D samples at each collection time point (**Figure 5C, Figure S3**). Among all timepoints, the *follow-up* collection (>9 months post-surgery) yielded the highest AUC (0.88, **Figure 5C, Figure S3**), indicating it was the most informative timepoint for assessing MRD. Based on this, the *follow-up* timepoint was selected for further analysis of the relationship between MRD status and patient survival (**Figure 5C**). We then established a threshold to classify MammaTrace-positive and −negative samples at *follow-up* using Youden’s index, which optimizes the combined sensitivity and specificity. At the resulting threshold of 0.897, 10/11 REC-D samples were MammaTrace-positive, corresponding to a sensitivity of 0.91 (**Figure 5D**, **Figure 6A**). While the AUC remains a valid measure of the classifier’s overall performance, sensitivity and specificity at the selected threshold may vary slightly in independent datasets due to sample-specific optimization. MammaTrace detected MRD in patients with liver, lung, bone, and brain metastases, among other metastatic sites, suggesting utility across a broad range of metastatic sites (**Table S9**). The one false negative patient (E100) had mediastinal and bone metastases (**Table S9**). Among DFS patients, a total of 33/40 were MammaTrace-negative, with a median post-surgical *follow-up* of 76 months, yielding a specificity of 0.83 (**Figure 5D**, **Figure 6A**). The one DN-IV sample was MammaTrace-positive at *follow-up*. In the REC-LR group, 1/3 tested positive (**Figure 5D**, **Figure 6A**). REC-LR samples were not used for ROC analysis or threshold determination.

**Figure 6:**
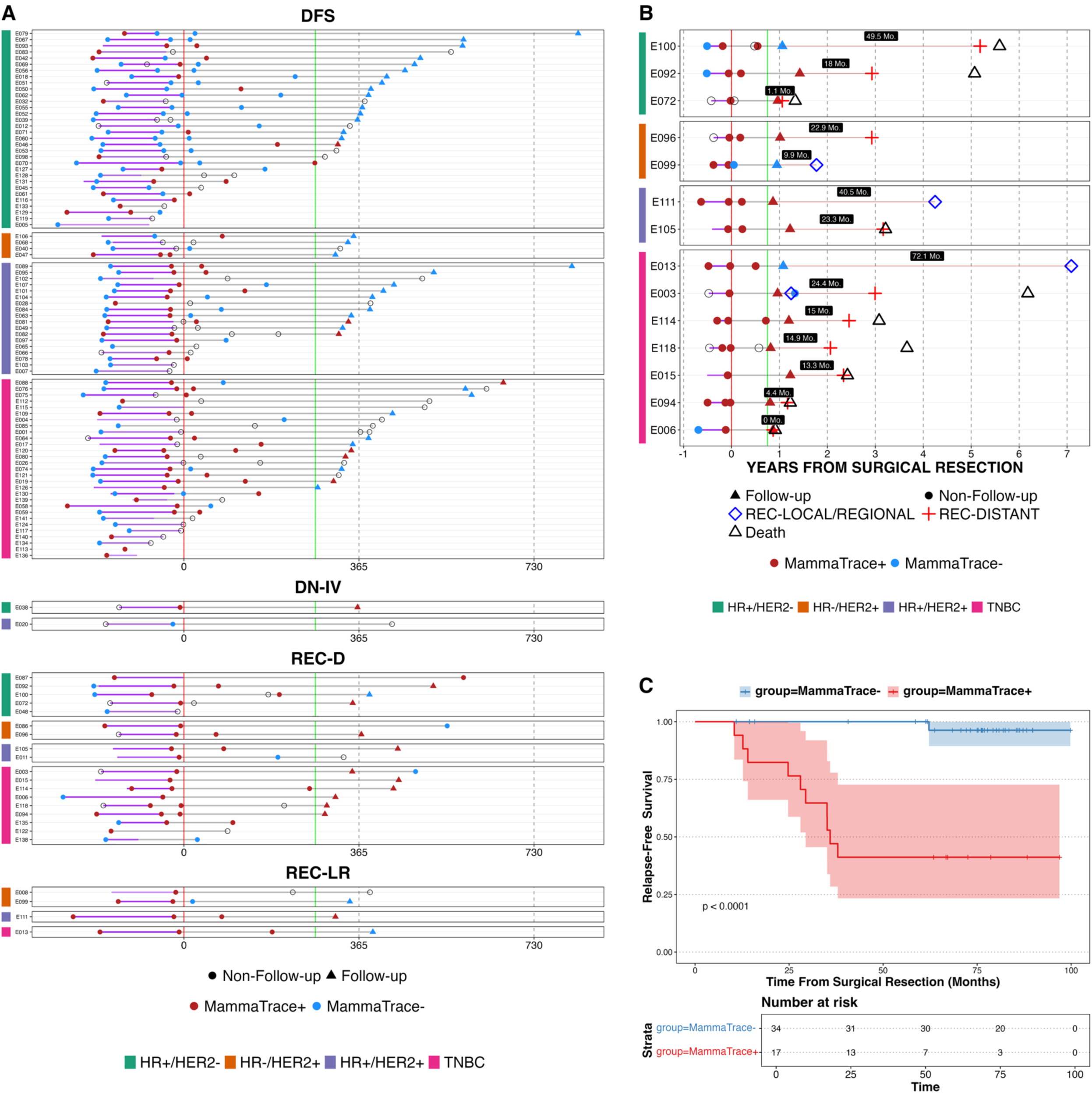
MammaTrace-positive results precede clinical recurrence. (**A**) Timeline of all cfDNA sample collections for each patient, colored by MammaTrace result (red = positive, blue = negative). Patients are grouped by disease status (DFS, DN-IV, REC-D, REC-LR), and molecular subtype is annotated on the left. (**B**) Samples from all REC-D and REC-LR patients with a follow-up blood collection (*N* = 14). Distant recurrence is marked with a red cross (*N* = 11), and locoregional recurrence with a blue diamond (*N* = 4); one patient (E003) experienced both. The lead time (in months) between a positive MammaTrace result and clinical recurrence is labeled for each patient. (**C**) Kaplan-Meier curve showing progression-free survival stratified by MammaTrace status at follow-up. Log-rank *p*-value and risk table are shown.

To investigate the reasons for the false positive results, we examined the 7 DFS patients that were MammaTrace positive at *follow-up*, but did not experience clinical recurrence. False positives were not associated with molecular subtype (2 HR+/HER2−, 1 HR+/HER2+, 4 TNBC), tumor stage (N stage I = 3, N stage II = 3, N stage III = 1), race or body mass index (**Figure S4**). All 7 patients were clinically disease-free at the study’s conclusion in July 2024, with a median disease-free survival of 6.04 years (range: 5.28–8.07 years). It remains possible that some of these patients may develop recurrence beyond the current follow-up period. Based on these findings, we determined that a *follow-up* interval of at least 9 months post-surgery was the most informative timepoint for MRD assessment using MammaTrace, as MammaTrace scores were most strongly correlated with subsequent distant recurrence.

### Lead time and prognostic value of MammaTrace for MRD+ patients

To assess the potential lead time for predicting clinical recurrence using MammaTrace, we analyzed the time interval between a Mamma-Trace positive result at *follow-up* and the date of clinically confirmed recurrence. Among the 10 MammaTrace-positive REC-D patients with *follow-up* collections, the median lead time was 15.0 months (457 days) from surgical resection (**Figure 6B**). Two patients had short lead times: one patient had a lead time of one day and a second patient had a lead time of one month (**Figure 6B**). All remaining patients had lead times ranging from 4 to 24.4 months, suggesting that MammaTrace could offer substantial early warning of recurrence. A MammaTrace-positive result at *follow-up* was also significantly associated with worse prognosis (**Figure 6C**) (log-rank p-value < 0.0001) and outperformed significantly better than pathologic complete response (**Figure S5**).

Taken together, these findings demonstrate that MammaTrace, using cfDNA methylation profiling, offers excellent performance for identifying patients with MRD at the end of definitive therapy who are at risk for future disease recurrence.

## DISCUSSION

The present study introduces MammaTrace, a tumor-naïve cfDNA methylation-based plasma-only liquid biopsy assay, specifically designed for MRD surveillance in breast cancer patients post curative-intent treatment. Informed by cfDNA methylation markers associated with MBC, MammaTrace demonstrated an excellent AUC of 0.88, effectively identifying early-stage patients likely to experience distant recurrence months to years before clinical or radiologic detection. For patients who developed MBC in our study, MammaTrace was positive in 10 of 11 cases at *follow-up*, with a median lead time of 15 months and up to 24 months. The single false-negative case had the longest disease-free interval (4.1 years) of any recurrent patient in the study, underscoring the importance of serial MRD testing. MammaTrace also detected a positive signal in one of the three patients who developed a locoregional recurrence, consistent with previous studies of reduced sensitivity in detecting local-only recurrences (19, 46). In patients who remained disease-free, MammaTrace correctly classified 33/40 as MRD-negative, with a median follow-up of 76 months. While seven patients were classified as false positive, all remained recurrence-free as of the study’s endpoint in July 2024, suggesting that some may still develop recurrence with longer follow-up. These findings highlight the potential of MammaTrace as an accurate tool for early detection of MRD and reinforce the need for ongoing, longitudinal monitoring in breast cancer survivors.

To our knowledge, MammaTrace represents the first tumor-naïve, cfDNA methylation-based, plasma only MRD surveillance assay developed specifically for breast cancer patients following curative-intent treatment. In contrast to tumor-informed MRD approaches, which rely on sequencing of primary tumor tissue, tumor-naïve assays like MammaTrace offer several advantages. Tumor-informed assays often require high-quality tumor tissue, which may not always be available or sufficient, and the process of panel development can take 6–8 or more weeks—delaying MRD detection and limiting the window for therapeutic intervention. While tumor-informed assays are highly specific, they may miss emerging or spatially heterogeneous disease due to tumor evolution or limited sampling. Tumor-naïve approaches circumvent these limitations, enabling faster implementation and broader applicability, particularly in real-world settings where tissue may be scarce (47). Our findings contribute to the growing evidence base for cfDNA-based MRD detection, offering a complementary or alternative approach to existing tumor-informed assays such as Signatera (Natera Inc.) and NeXT (Personalis Inc.). Unlike tumor-informed methods that rely on tissue biopsies and are constrained by tissue availability and tumor purity, MammaTrace uses a plasma-only tumor-naïve approach designed to overcome these limitations. The strategy, however, is partially informed by leveraging data from cfDNA methylation patterns linked to metastasis across different breast cancer subtypes and racial backgrounds, such as from AA patients with TNBC. Conducting feature discovery directly in plasma, rather than relying on primary EBC tissue or metastatic tumor tissue—which is often unavailable, heterogeneous, or derived from different organ sites—offers a major clinical advantage. This approach makes cfDNA methylation profiling more practical and scalable for real-world MRD implementation, particularly in metastatic settings. cfDNA also reflects not only tumor-derived DNA but also systemic factors, capturing a broader, clinically relevant signal that may be absent in tumor tissue. This enhances the assay’s ability to detect residual disease across various stages of cancer progression by identifying methylation changes that are commonly linked to the metastatic potential of breast cancer cells, without the need for prior tumor-specific profiling and offers a more accurate, less complex and more broadly applicable solution for MRD detection. The Signatera MRD assay (Natera Inc.) (18, 19, 20, 21, 45, 48, 49) has been evaluated in multiple EBC studies (19, 38). In the initial landmark study, Coombs et al. found that Signatera was able to detect breast cancer MRD with 89% sensitivity and 100% specificity with a median lead time of 8.9 months (19). A second EBC Signatera study showed similar results in a larger EBC cohort (sensitivity = 88%, specificity = 95%, median lead time = 10.5 months) (38). In the TRACERx study, Abbosh et al. found that the NeXT assay (Personalis Inc.) predicted lung cancer recurrence with 49% sensitivity and 96% specificity at a 4-month landmark timepoint (50). While these early findings are promising, both MRD assays are tumor-informed and require primary tumor samples with high tumor purity to maintain sensitivity (51), as well as the creation of bespoke multiplex panels which add logistical complexity and cost.

Moreover, tumor-naïve plasma-only approaches are not only lower in cost, complexity, and turnaround time (18, 52), but offer a strategy for MRD detection in situations where sufficient tumor tissue is not available, which is often the case after treatment (53). Recent work by Parikh et al. found that a tumor-naïve assay using plasma-only circulating DNA methylation and genomic alterations as markers (Reveal, Guardant Inc.), demonstrated comparable sensitivity and specificity to tumor-informed approaches in colorectal cancer patients (18, 48). This assay was also used to predict breast cancer recurrence in early-stage patients and was found to have 80% sensitivity and 100% specificity in predicting distant recurrence (N = 5) (46).

The longitudinal design of our study enabled us to assess the prognostic value of MammaTrace at multiple timepoints throughout treatment, and to define the *follow-up* timepoint for analysis based on performance. Interestingly, after neoadjuvant treatment (*post-neo*) and surgical resection (*post-op*), we observed an increase in the MammaTrace score (cfDNA methylation) in some patients. This finding contrasts with those from somatic variant-based, tumor-informed ctDNA (mutation-based) MRD assays, which typically show clearance of ctDNA mutations during and after chemotherapy (40, 41, 42, 43, 44, 45). This observation suggests that methylation-based assays may capture distinct biological processes that are not reflected in mutation-based ctDNA assays. The reason for the increase in the MammaTrace scores following neoadjuvant chemotherapy is not fully understood. While ctDNA mutation clearance typically reflects a reduction in tumor burden, elevated MammaTrace scores may indicate biological changes such as cellular stress responses, host tissue remodeling, or the emergence of resistant cell populations (54, 55). These treatment-resistant persisting cells may undergo epigenetic alterations that facilitate adaptation to chemotherapy. Thus, an increase in MammaTrace score in post-treatment plasma may reflect changes in DNA methylation that accompany tumor cytoreduction as well as the development of resistant clones. Importantly, patients whose disease recurred had sustained high MammaTrace scores compared with disease-free patients who became MRD-by the defined *follow-up* timepoint. This indicates that clearance of altered, methylated DNA in response to chemotherapy, that likely arises from affected tissues in addition to the cancer, does clear, and that tumor-specific methylated DNA persists, indicating MRD.

An important aspect of this study is the inclusion of cfDNA methylation features across all major breast cancer subtypes in the development of the MammaTrace assay. Our analysis revealed that methylation profiles varied significantly among the different molecular subtypes, consistent with prior research (56, 57, 58, 59). Notably, this is the first study to identify subtype-specific methylation alterations detectable in cfDNA and to integrate these DMRs into a cfDNA-based methylation MRD assay that is applicable across breast cancer molecular subtypes.

Few studies on MRD in solid tumors specifically address racial differences or include patients with diverse racial backgrounds in their machine learning model building. Most published MRD studies have focused on predominantly white populations, potentially overlooking genetic and epigenetic variations that could influence assay sensitivity and specificity across different racial groups. MammaTrace explicitly incorporated methylation features of cfDNA from AA patients with TNBC as well as normal cfDNA from AA healthy controls, in addition to including normal cfDNA samples from various other racial/ethnic backgrounds. Interestingly, our analysis revealed that cfDNA methylation patterns differ between AA and non-AA TNBC patients, highlighting the importance of building models that capture different populations. MammaTrace performed equally well in detecting MRD in both AA and non-AA TNBC patients because the model was trained on cfDNA methylation from AA TNBC and Non-AA TNBC patients. This approach not only enhances the inclusivity of MRD surveillance but also addresses potential variations in cancer outcomes related to genetic and epigenetic differences across races that need to be considered.

The study has some limitations that warrant consideration. While the overall cohort was sufficient to demonstrate statistical significance and robust classifier performance, the sample size in certain subgroups was relatively small, and some cfDNA samples were not evaluable due to limited material—reducing the number of patients with complete longitudinal timepoints. This may limit broader generalizability and underscores the need for further validation in larger cohorts. In addition, the study included only a single follow-up collection per patient, which limited our ability to assess dynamic changes in MRD status over the course of long-term survivorship—such as whether MRD-negative patients later become positive. This is particularly important in breast cancer, where recurrence can occur many years after initial treatment. Finally, although MammaTrace was trained using data from patients of diverse racial backgrounds, additional validation in larger, multi-ethnic cohorts is necessary to confirm performance across populations and explore any potential race-specific biases.

In conclusion, the MammaTrace cfDNA assay shows substantial promise as a non-invasive, tumor-naïve plasma-only method for MRD surveillance in breast cancer. Its ability to predict recurrence with a median lead time of 15 months highlights its potential to enable early intervention strategies. Future studies should aim to expand the diversity of patient cohorts, further optimize sensitivity and specificity, and explore the assay’s utility in guiding treatment decisions in clinical practice.

## DECLARATIONS

### Data availability

Some data underlying this study are not publicly available at this time due to ongoing intellectual property considerations related to a licensed patent. This is in accordance with NIH guidelines, which permit data-sharing exceptions under specific circumstances, including the protection of proprietary information. We are committed to responsible data stewardship and to complying fully with NIH policies, and we welcome reasonable requests for data access that are consistent with our licensing obligations and ongoing translational efforts.

### Authors’ contributions

Study conception, BS; experimental design, BS, BP, DB, GG; data generation, BS, DB, AK, GG, MP, JG, BP; data analysis, BS, DB, JPL; manuscript preparation, DB, AK, GG, JPL, DS, JC, DS, HJL, CHH, CL, JO, BP. All authors read and approved the final manuscript.

## Acknowledgements

Not applicable

---

**<u>Figure S1: Correlation matrix of control samples across sequencing runs.</u>** The figure displays beta value correlations for two control samples sequenced across seven independent runs. Scatterplots show the spread of beta values between sequencing runs. Correlation coefficients and p-values are indicated in the upper portion of the plot. Histograms show the distribution of beta values from 0 to 1.

**<u>Figure S2: Paired MammaTrace scores by patient across treatment timepoints.</u>** Each panel shows the change in MammaTrace score for paired samples collected at two timepoints, stratified by disease status (DFS, REC-D, and REC-LR). Each point represents an individual patient, with connecting lines indicating the direction of change (increase or decrease) in MammaTrace score. Boxplots summarize the overall score changes between collection timepoints. Wilcoxon’s signed rank p-values are indicated at the top of each plot – the total number of patients in each plot is indicated in the bottom right.

**<u>Figure S3: ROC analysis at each timepoint.</u>** ROC curves show the performance of MammaTrace at each collection timepoint. AUC and number of patient samples in each analysis is indicated.

**<u>Figure S4: MammaTrace scores by patient characteristics</u>**. The figure shows MammaTrace scores for DFS patients by subtype (**A**), stage (**B**), race (**C**) and BMI (**D**). False positive status is indicated by red dots.

**<u>Figure S5: Kaplan-Meier curves of relapse-free survival stratified by pathologic complete response (pCR</u>).** Kaplan-Meier curves compare relapse-free survival between patients who achieved pCR and those with residual disease following neoadjuvant chemotherapy. **(A)** Analysis limited to patients included in the follow-up timepoint (same as in Figure 6C). **(B)** Analysis including all patients in the study cohort.

**<u>Table S1:</u>** Summary of all MBC, healthy and EBC cfDNA plasma samples received for this study.

**<u>Table S2:</u>** Summary of all MBC and healthy patients included in this study.

**<u>Table S3:</u>** Summary of all EBC patients from whom cfDNA samples were included in this study.

**<u>Table S4:</u>** Tumor stage information for AA TNBC patients included in the study.

**<u>Table S5:</u>** Summary of EBC samples analyzed using MammaTrace hybrid probe capture, by disease status and collection timepoint.

**<u>Table S6:</u>** Number of unique patients contributing to each MBC or healthy cfDNA pool.

**<u>Table S7:</u>** Treatment-naïve EBC samples used for WGBS feature discovery.

**<u>Table S8:</u>** Summary of MBC and healthy samples used for MammaTrace validation and model construction.

**<u>Table S9:</u>** Recurrence site information for the 11 REC-D patients included in the follow-up timepoint analysis.

