## Supplementary figures and images for "MammaTrace – a cell-free DNA methylation plasma only assay for minimal residual disease detection in breast cancer patients"

### Supplemental Figures

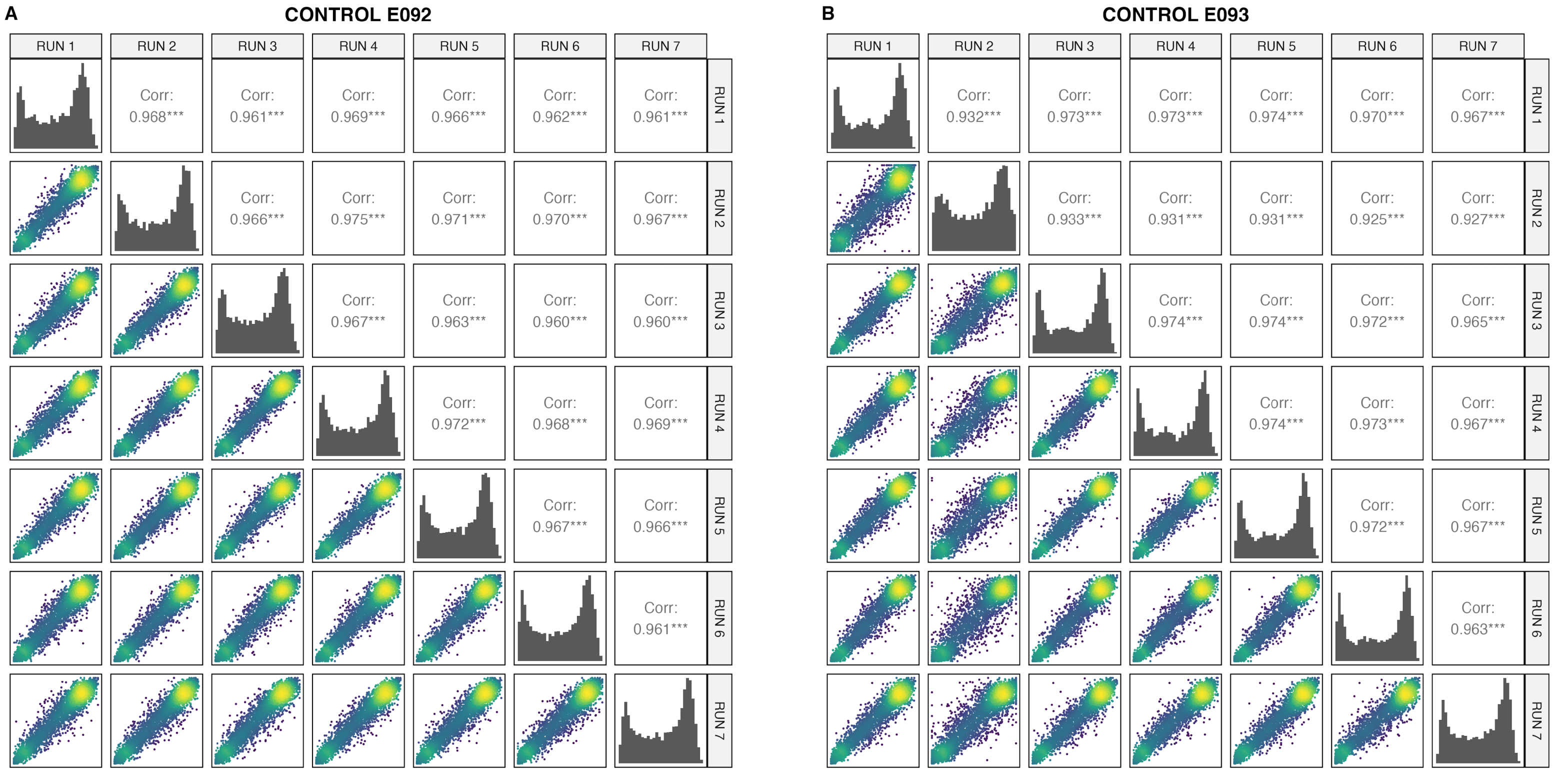

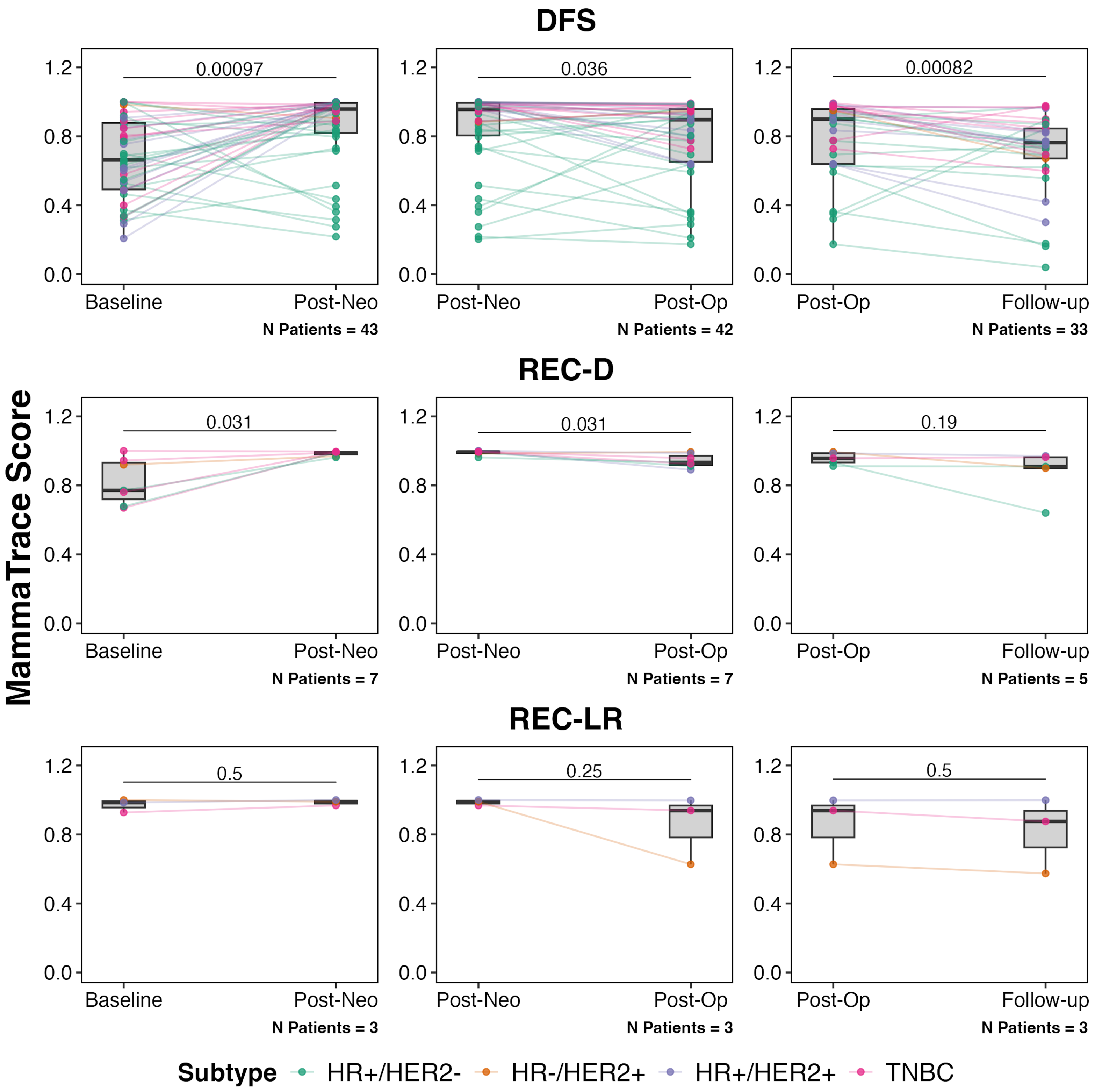

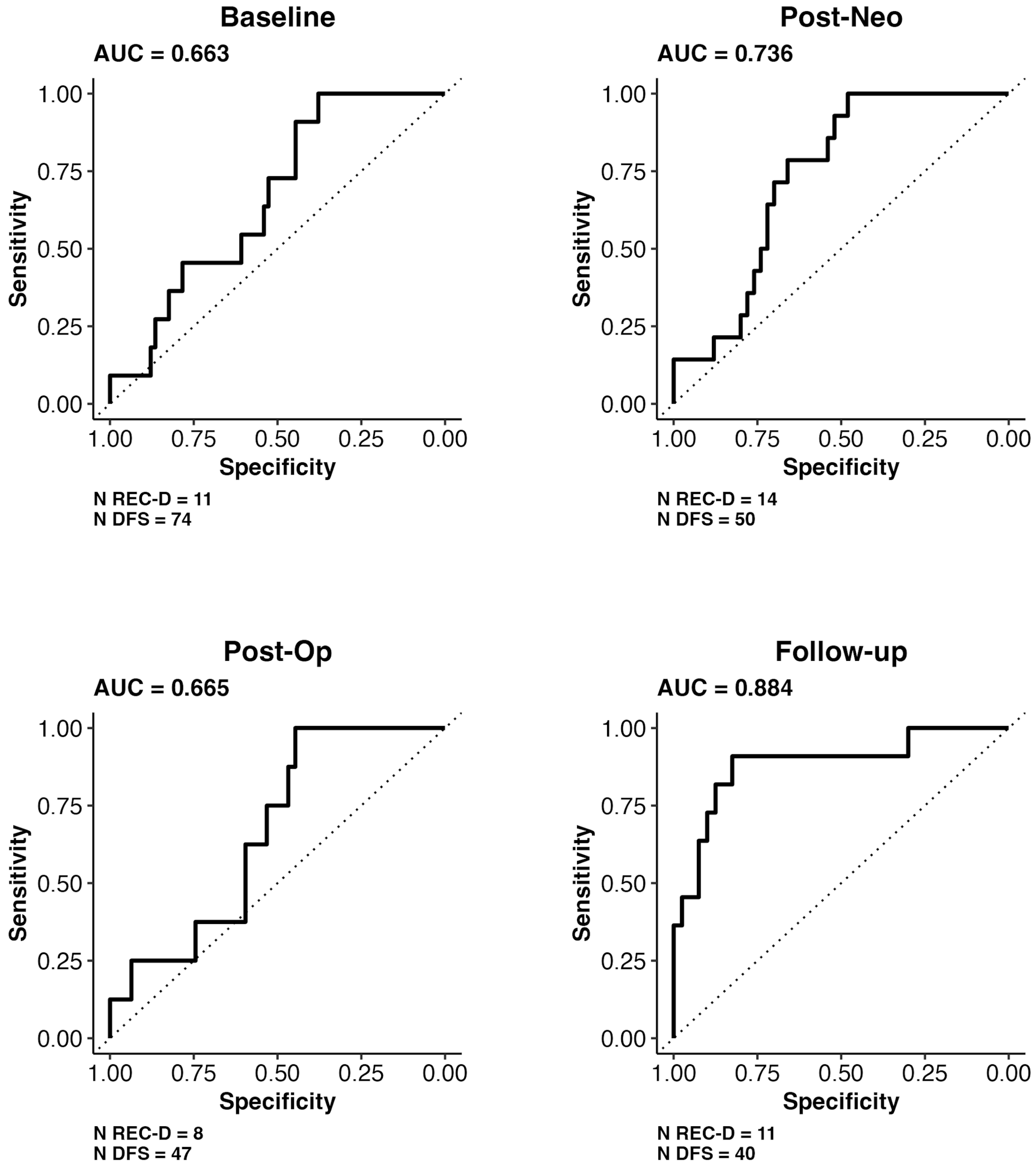

#### Figure S4

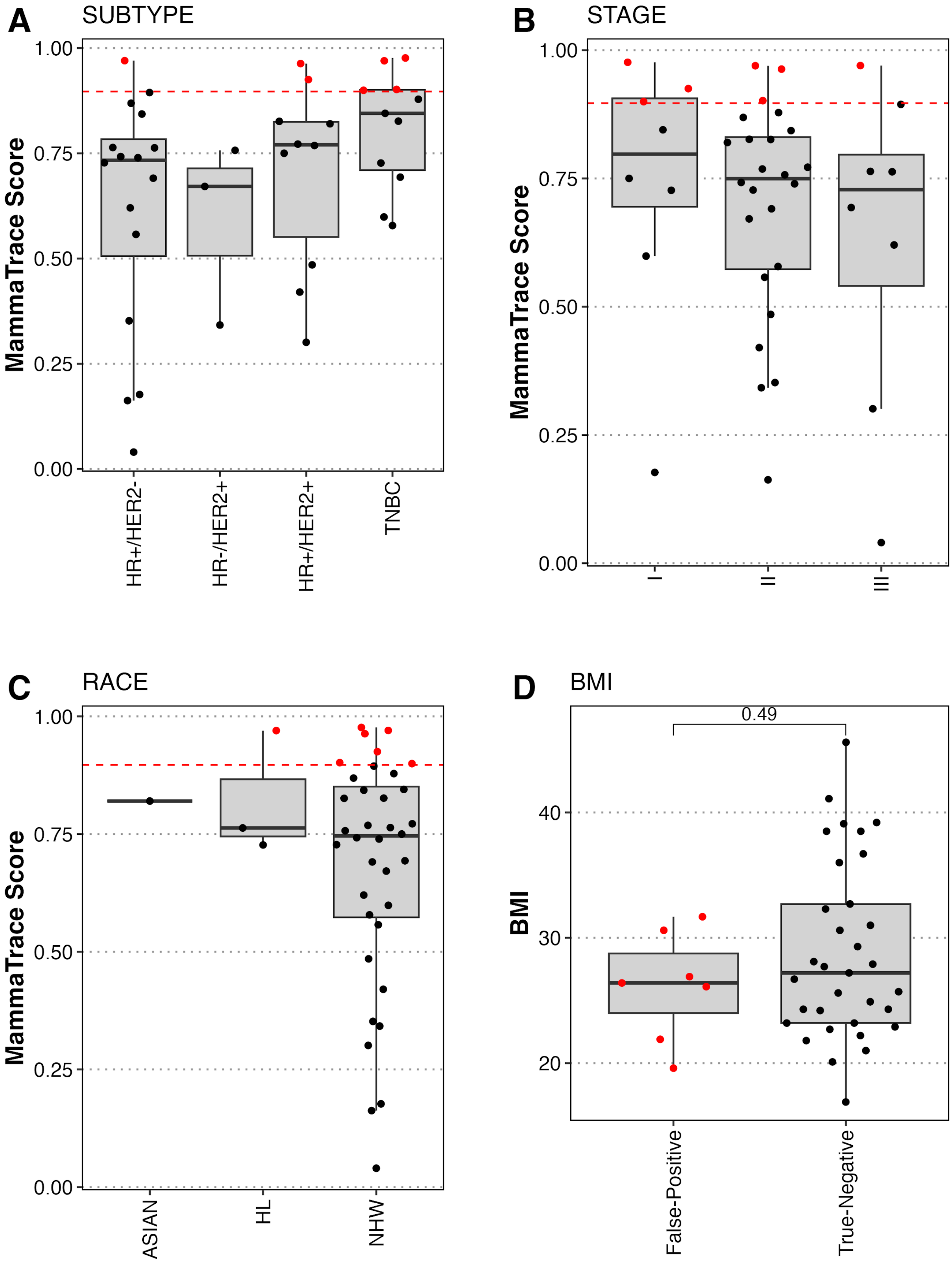


● False-Positive ● True-Negative

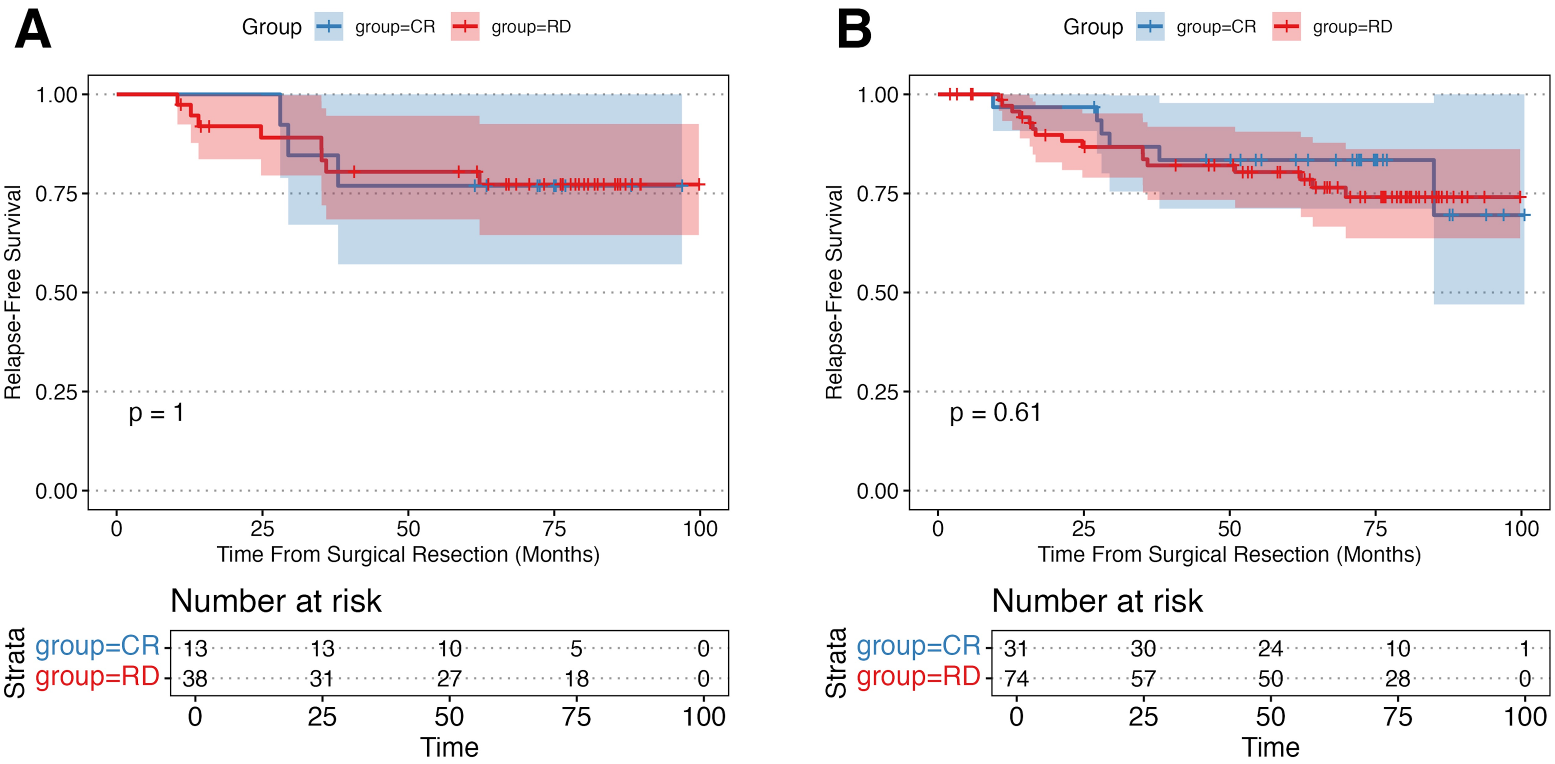
